# Potential diagnostic biomarkers for human mesenchymal tumors, especially LMP2/β1i and cyclin E1 differential expression

**DOI:** 10.1101/2024.06.27.24309614

**Authors:** Yuriko Tanabe, Takuma Hayashi, Okada Mako, Hiroyuki Aburatani, Kaoru Abiko, Ikuo Konishi

## Abstract

**Objectives:** Most mesenchymal tumors found in the uterine corpus are benign tumors; however, uterine leiomyosarcoma is a malignant tumor with unknown risk factors that repeatedly recurs and metastasizes. In some cases, the histopathologic findings of uterine leiomyoma and uterine leiomyosarcoma are similar and surgical pathological diagnosis using excised tissue samples is difficult. It is necessary to analyze the risk factors for human uterine leiomyosarcoma and establish diagnostic biomarkers and treatments. Female mice deficient in the proteasome subunit low molecular mass peptide 2 (LMP2)/β1i develop uterine leiomyosarcoma spontaneously.

**Methodology:** Out of 334 patients with suspected uterine mesenchymal tumors, patients diagnosed with smooth muscle tumors of the uterus were selected from the pathological file. To investigate the expression status of biomarker candidate factors, immunohistochemical staining was performed with antibodies of biomarker candidate factors on thin-cut slides of human uterine leiomyosarcoma, uterine leiomyoma, and other uterine mesenchymal tumors.

**Results:** In human uterine leiomyosarcoma, there was a loss of LMP2/β1i expression and enhanced cyclin E1 and Ki-67 expression. In human uterine leiomyomas and normal uterine smooth muscle layers, enhanced LMP2/β1i expression and the disappearance of the expression of E1 and Ki-67 were noted. The pattern of expression of each factor in other uterine mesenchymal tumors was different from that of uterine leiomyosarcoma.

**Conclusions:** LMP2/β1i, cyclin E1, and Ki-67 may be candidate factors for biomarkers of human uterine leiomyosarcoma. Further large-cohort clinical trials should be conducted to establish treatments and diagnostics for uterine mesenchymal tumors.

## Introduction

The uterus, the organ that harbors embryos and fetuses, has the following three layers: the endometrium, which acts as a bed for the embryo, the uterine smooth muscle layer (myometrium) that protects the embryo, and the serous membrane (perimetrium) that envelops the uterus. In general, endometrial tumors represent epithelial malignancies of the endometrium and are broadly classified into tumors that develop in the cervix or inside the uterus. If smear screening for cervical cancer confirms the onset of cervical cancer early enough, the rate of cervical cancer-related mortality would decrease. Conversely, the incidence of endometrial cancer is increasing because simple methods of screening for endometrial cancer have not been established. Most cervical tumors are malignant tumors classified as squamous cell carcinoma and adenocarcinoma. Mesenchymal tumors that develop in the uterine smooth muscle layer are conventionally classified into benign uterine leiomyoma (uLMA) and malignant uterine leiomyosarcoma (Ut-LMS) based on cytological atypia, mitotic activity, and other criteria. Ut-LMS is relatively rare, with an estimated annual incidence of 0.64 per 100,000 women (1). Ut-LMS occurs more frequently in the smooth muscle layer of the uterine body than in the cervix. Because Ut-LMS is resistant to chemotherapy and radiotherapy, surgery is virtually the only viable treatment option (2). The prognosis of Ut-LMS is poor, with a 5-year overall survival rate of about 20% (3, 4). However, the development of efficient adjuvant therapies for Ut-LMS is expected to improve the prognosis of patients with Ut-LMS. The prevalence of uLMA in women aged up to 50 years is 70%∼80% (FACT SHEET - Uterine Fibroids, 2010). The diagnosis of uterine mesenchymal tumors is assigned cases (5,6,7) using the diagnostic category and morphological criteria for uterine mesenchymal tumors (Note 1). However, due to the similarity in cell morphology of uLMA and Ut-LMS, it is hard to differentiate between uLMA and Ut-LMS in some cases. Therefore, the surgical pathological diagnosis is made by macroscopic and histopathological findings on excised tissue specimens (5). Since nonstandard subtypes of uterine mesenchymal tumors such as endometrial stromal sarcoma and liposarcoma are classified based on the characteristics of these tumor cells, it is important to establish a diagnostic method for nonstandard smooth muscle cell differentiation in actual clinical practice.

Much remains unknown about the molecular mechanisms involved in the pathogenesis of uLMA and Ut-LMS. Tumors that develop in the myometrium for any reason are thought to gradually grow in size and form tumors due to the effects of somatic mutations by female hormones or mediator complex subunit 12 (MED12) (8). However, to the best of our knowledge, the involvement of female hormones in the pathogenesis of Ut-LMS has not been observed and no obvious risk factors have been found. Cases of Ut-LMS with hypocalcemia or eosinophilia have been reported; however, neither clinical abnormality is an initial risk factor for Ut-LMS. The identification of risk factors associated with the development of Ut-LMS contributes to the development of preventive measures and novel treatments.

Cellular proteins are predominantly degraded by a protease complex consisting of 28 subunits of 20–30 kDa, which is called the 20S proteasome (9). The proteasome degradation pathway is essential for many cellular processes, including the cell cycle, gene expression regulation, and immune functions. Interferon-γ (IFN-γ) induces the combinational expression of the proteasome subunit’s low-molecular-weight polypeptides (LMP)2/β1i, LMP7/β5i, and LMP10/β10i. Molecular approaches to studying the correlation between IFN-γ and tumor cell proliferation have attracted much attention (10). LMP2/β1i expression may be associated with cell survival (11, 12). Homozygous mice deficient in LMP2/β1i show abnormalities in tissue-dependent or substrate-dependent proteasome biological functions. Ut-LMS occurred in female LMP2/β1i-deficient mice aged ≥6 months, with an incidence of uterine leiomyosarcoma of approximately 40% by 14 months of age (13, 14). The curve of the incidence showing the prevalence of Ut-LMS in mice by month is similar to the curve showing the morbidity of human Ut-LMS that occurs after menopause.

Advances in cell cycle research have demonstrated that interactions between cyclins, cyclin-dependent kinases (CDKs), and tumor suppressor gene products play an essential role in cell cycle progression (15). Cyclins, which form complexes with CDKs, are a group of proteins that are periodically expressed during the cell cycle (16). The expression of these cyclins, CDKs, or cell cycle regulators is regulated by proteasome activation. Thus, compared with normal uterine smooth muscle cells, different expression levels of these cell cycle-related factors are observed in uterine leiomyosarcoma cells that have lost LMP2/β1i expression (17). Per previous experimental results, cyclin E1 is overexpressed in human mesenchymal tumors (18, 19, 20, 21). Therefore, an expression vector of LMP2/β1i can be inserted into uterine leiomyosarcoma cells (SK-LMS) that have lost LMP2/β1i expression, and LMP2/β1i stable transformant SK-LMS cells were created in which LMP2/β1i is constitutively expressed. To investigate the differences in gene expression between SK-LMS cells and stable transformant SK-LMS cells, gene profiling was created using these two cell types. We, the medical staff, used the factors specifically expressed in Ut-LMS cells obtained from the results of gene profiling as specific marker candidate factors for uterine leiomyosarcoma cells. In this clinical study, to develop targeted therapeutics and diagnostic methods for Ut-LMS, we investigated the expression levels of Ut-LMS cell-specific marker candidate factors in extracted tissues obtained from patients suspected of developing uterine mesenchymal tumors by the immunohistochemical (IHC) staining using the appropriate monoclonal antibodies and our results were examined. Based on the results of the clinical study, we verified the biomarker candidate factors for Ut-LMS. Five candidate factors were identified as pathological variants of Ut-LMS, including LMP2/β1i and cyclin E1. These biomarker candidate factors have the potential to be diagnostic biomarkers and target molecules for new therapeutic approaches.

## Materials and methods

### Tissue collection

A total of 334 patients aged 32–83 years with suspected development of uterine smooth muscle tumors were selected from the histopathological file (22, 23). Continuous sections were taken from at least two tissue blocks in each patient for hematoxylin and eosin staining and IHC. We performed an analysis of the removed tissue after obtaining each patient’s written consent. This clinical study was approved by the ethics committees of Shinshu University, Kyoto Medical Center, and Kyoto University.

### Immunohistochemistry (IHC) experiment

To examine the expression levels of the five candidate factors, IHC staining was performed on sequential sections of excised tissue from uterine mesenchymal tumors containing human Ut-LMS or uLMA. The anti-human cyclin E1 monoclonal antibody was purchased from Immunotech (Marseille, France). The anti-human LMP2/β1i antibody was produced in a joint research and development project of SIGMA-Aldrich Israel Ltd. (Rehovot, Israel) and our research team. Anti-human cyclin B monoclonal antibodies and anti-human caveolin monoclonal antibodies were purchased from Santa Cruz Biotechnology, Inc. (Santa Cruz, CA, USA). The IHC experiment was performed using the avidin-biotin complex method described above. Briefly, we prepared a 5-mm-thick slice of tissue sections from paraffin-embedded hysterectomy samples from each patient with a mesenchymal tumor. Tissue sections from the patients were deparaffinized and rehydrated in graded concentrations of ethyl alcohol. Tissue sections were incubated with normal mouse serum for 20 minutes and then incubated with primary antibodies for 4 hours at room temperature, after which they were incubated with a biotinylated secondary antibody (Dako, CA) and reacted with a streptavidin complex (Dako). Reacted tissue sections were stained with a solution of 3,3-diaminobenzidine (DAB; Dako, CA) and counterstained with hematoxylin. The normal myometrial portion of the tissue section as a specimen was used as a positive control. The negative control consisted of tissue sections incubated with normal rabbit IgG instead of the primary antibody. These experiments were conducted at Shinshu University, the National Hospital Organization Kyoto Medical Center, and Kyoto University Hospital per local guidelines (Approval No. M192).

## Result

Mice deficient in the *Lmp2/β1i* gene have been reported to develop Ut-LMS spontaneously [24]. Compared with normal uterine smooth muscle and uterine smooth muscle type tissues, LMP2/β1i expression is significantly lower or negative in Ut-LMS tissues. Therefore, defective LMP2/β1i expression is considered one of the risk factors for Ut-LMS [25118]. The antitumor response mediated by MHC class I molecules is a process influenced by the function of the proteasome induced by stimulation with interferon γ (IFN-γ) [26,27]. These findings support the fact that IFN-γ prevents the development of primary tumors and exhibits tumor suppression effects through immune responses [26,28]. LMP2/β1i expression is significantly induced by IFN-γ, and the activation of signal transducer and activator of transcription (STAT) 1 by stimulation with IFN-γ induces upregulation of tumor suppressors, including interferon regulatory factor 1 (IRF1). IRF1 functions as a transcriptional regulator that plays an important role in the regulation of LMP2/β1i expression [29, 30]. Reduced IRF1 levels caused by LMP2/β1i deficiency could constitute a risk factor for Ut-LMS [25]. In 85% of tissues removed from patients with Ut-LMS, a marked decrease in LMP2/β1i protein expression is noted. The results of LMP2/β1i immunostaining are useful for the differential diagnosis of Ut-LMS and ULMA [31]. LMP2/β1i is a promising diagnostic marker candidate factor for Ut-LMS, and it has the potential to become a target molecule for new therapies. One Ut-LMS case was stained with LMP2/β1i.

Since the expression of cyclin E1 in Ut-LMS was observed to be higher than that of uLMA and normal uterine smooth muscle layers, this clinical study aimed to verify the expression of cyclin E1 in the extracted tissue obtained from each case. The results of IHC experiments showed that staining with cyclin E1 was observed in all tissue samples obtained from patients with Ut-LMS. Pathological examinations of excised tissue samples revealed the presence of a mass with a diameter of up to 3 cm in the lumbar quadriceps muscle without a fibrous capsule. All lymph node tissues tested negative for Ut-LMS metastases. The results of the IHC experiments showed the positive expression of cyclin E1 and Ki-67 and the negative expression of LMP2/β1i. The histopathologic findings of metastatic Ut-LMS of skeletal muscle and rectal lesions were consistent with the primary histopathologic findings of Ut-LMS.

Our medical staff has previously demonstrated an association between the abnormal expression of the female hormone estrogen receptor and tumor protein 53 (TP53) and the proliferation marker Ki-67 (Ki-67) and the mutation of TP53 and the pathogenesis of Ut-LMS. LMP2/β1i expression deficiency is associated with these factors (32,33). Compared with these factors, LMP2/β1i expression deficiency is highly useful as a feature of Ut-LMS (Table). In subsequent experiments, both estrogen receptor and progesterone receptor were stained in almost all uLMAs, regardless of the phase of the menstrual cycle, and the number of Ki-67 positive cells in the uLMA was significantly lower than in the Ut-LMS (Table). In addition, in IHC staining, almost all uLMAs showed TP53 staining (Table). The IHC experiment performed using candidate biomarkers of uterine mesenchymal tumors (i.e., cyclin E, Ki-67, LMP2/β1i) may facilitate the differentiation of smooth muscle tumor uncertain malignancy potential (STUMP), which is difficult to diagnose via histopathological examinations of excised tissue samples, and uterine leiomyomas and leiomyosarcoma.

STUMP is a general term for uterine mesenchymal tumors whose malignancy is difficult to determine through pathological examination based on the malignancy category and findings such as cytoskeleton. In a clinical study conducted by our research group, IHC testing was performed using anti-human cyclin E monoclonal antibody and anti-human Ki-67 monoclonal antibody on tumor tissues removed by surgical treatment from patients who were diagnosed with Ut-LMS or STUMP and died within 5 years after surgical treatment (Ut-LMS = 10 cases, STUMP = 7 cases). Compared to Ut-LMS tumor tissue, the percentage of Ki-67 positive cells in STUMP tumor tissue is lower (Figure 1). However, the expression level of Ki-67 in STUMP tumor tissues was higher than that in Ut-LMS tumor tissues (Figure 1). Compared to Ut-LMS tumor tissue, the percentage of cyclin E-positive cells in STUMP tumor tissue is slightly lower (Figure 1). However, the expression level of cyclin E in STUMP tumor tissues was higher than that in Ut-LMS tumor tissues (Figure 1). These results suggest that cases of gynecological mesenchymal tumors whose malignancy was not determined by surgical pathology and who were differentiated as STUMP and whose IHC tests showed high expression levels of cyclin E and ki-67 may have a poor prognosis. The expression levels of cyclin E and ki-67 in uterine mesenchymal tumors are considered to be candidate factors for predictive markers of life prognosis.

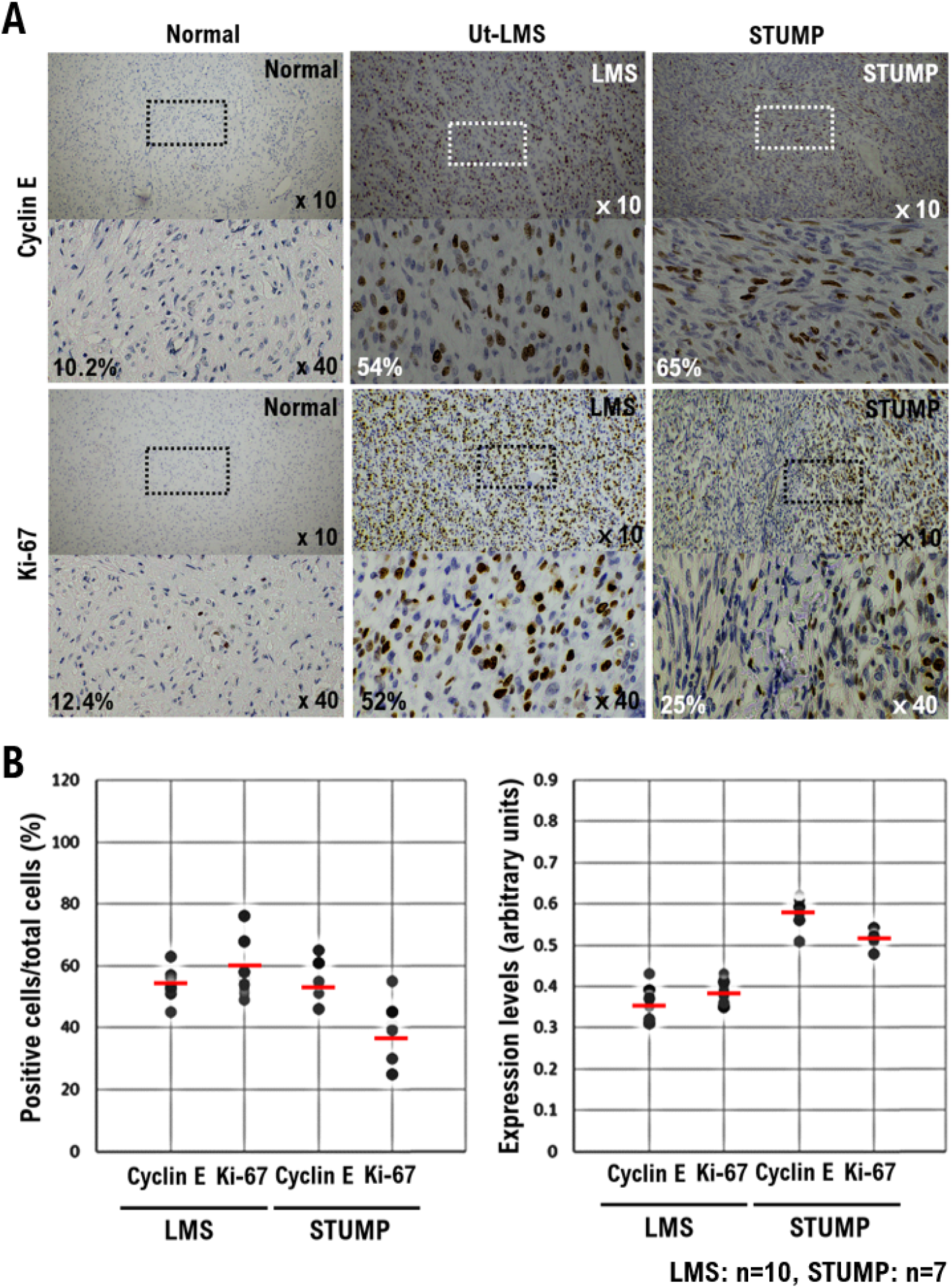

## Discussion

Recent clinical reports have shown the expression of Lmp2/β1i mRNA and proteins in luminal and glandular epithelium, chorionic placenta, chorionic pancreatic cells, and arterial endothelial cells (34). These results suggest that LMP2/β1i is involved in placental villus infiltration, extracellular matrix degradation, immune tolerance, glandular secretion, and angiogenesis (34). This study is expected to help elucidate the regulatory role of LMP2/β1i in embryo implantation. Cyclin E1 immunoreactivity was observed in the nucleus and cytoplasm of cells in all Ut-LMS cases studied, while most cases of uLMA and normal myometrium tested negative for cyclin E1. Cyclin E1 is a regulatory protein involved in mitosis, and CDK and the gene product complex combine to form a maturation-promoting factor (MPF) (35). Cyclin E1/CDK2 is involved in early mitotic events such as chromosomal condensation, nuclear membrane disruption, and spindle pole assembly. When cyclin E1 levels are depleted, the cyclin E1/CDK1 complex cannot be formed, and cells cannot enter the M phase, which slows down cell division. Some anticancer therapies are designed to prevent the formation of cyclin E1/CDK2 complexes in cancer cells and slow down or prevent cell division (36). While most of these methods target the CDK2 subunit, there is growing interest in targeting cyclin E1 as well in the field of oncology. In other words, cyclin E1 regulation may provide clues to the development of new treatment options for uterine leiomyosarcoma. However, clinical risk factors for its development have not been identified due to the absence of suitable experimental animals. LMP2/b1i-deficient mice were the first spontaneous animal models of Ut-LMS. Therefore, poor LMP2/b1i expression might be one of the causes of Ut-LMS.

To date, several candidate factors as biomarkers have been reported for uterine leiomyosarcoma. For example, 5’-Nucleotidase Domain Containing 2 (NT5DC2) and tyrosine kinase-like orphan receptor 1 (ROR1) have been identified as possible factors that induce Ut-LMS proliferation. It was not detected as a biomarker candidate for Ut-LMS (37, 38). In actual clinical practice, cancer genomic medicine is used to select new antitumor agents for patients with advanced recurrent cancer by cancer gene panel testing. To date, new antitumor agents have been selected for several patients with advanced recurrent Ut-LMS by oncology gene panel testing; however, pathological variants of NT5DC2 and ROR1 molecules have not been detected.

Nevertheless, this clinical study has a few limitations, the main one being the fact that it was conducted in a cohort with a small number of cases. Therefore, to adequately demonstrate whether LMP2/β1i and cyclin E1 are potential biomarkers to differentiate between Ut-LMSs and uLMAs, the reliability and properties of LMP2/b1i and cyclin E1 as diagnostic indicators are being investigated at several clinical research sites. Clinical research is not over yet, and large-scale clinical trials need to be conducted to verify the findings obtained this time.

## Conclusion

The correlation between candidate biomarkers for uterine mesenchymal tumors (LMP2/β1i, cyclin E1, Ki-67) and the development of Ut-LMS has been revealed, and the identification of specific risk factors may lead to the development of new treatments for this disease. Ut-LMS is resistant to chemotherapy and has a poor prognosis. Molecular biological and cytological information derived from further research experiments with human tissues and LMP2/β1i-deficient mice will contribute significantly to the development of prophylaxis, potential diagnostic biomarkers, and new therapies for Ut-LMS.

(Note 1) The typical macroscopic appearance is a large (>10 cm), poorly bounded mass with a soft, fleshy consistency and a gray-yellow to pink variegated cut surface with foci of hemorrhage and necrosis (6,7). The histological classification of uterine sarcoma is based on homology with normal cell types and includes human Ut-LMS (similar to the myometrium), stromal sarcoma (similar to the endometrial stroma), and other heterogeneous cell types (i.e., osteosarcoma and liposarcoma). Microscopically, most human Ut-LMS are clearly malignant, with hypercellularity, coagulative tumor cell necrosis, and abundant mitoses [>10–20 mitotic figures per 10 high-power fields] mf)], atypical mitosis, and cytological atypia. The mitotic rate is the most important determinant of malignant tumors; however, it is corrected by the presence of necrosis and cytological atypia. The diagnosis of human Ut-LMS can be made in the presence of tumor necrosis and mitosis. In the absence of tumor necrosis, the diagnosis can be made in the case of moderate-to-severe cytological atypia and a mitotic index greater than 10 mf/10 hpf. In the absence of necrosis and significant atypia, a high mitotic index is compatible with a benign clinical course; however, supporting data are limited (6,7).

## Data Availability

All data produced in the present study are available upon reasonable request to the authors

## Acknowledgments

I would like to express my sincere gratitude to Prof. Luc Van Kaer (Vanderbilt University Medical Center) for their support of the research experiment. This research was supported in part by the Ministry of Education, Culture, Sports, Science and Technology, the Osaka Cancer Research Foundation, the Ichiro Kanehara Foundation for the Promotion of Medicine and Medical Care, the Cancer Research Foundation, the Kanazawa Medical Research Foundation, the Shinshu Medical Foundation, and the Takeda Pharmaceutical Science Foundation.

## Data Availability

The authors declare that data supporting the findings of this study are available within the article.

## Ethics approval and consent to participate

This study was reviewed and approved by the Central Ethics Review Board of the National Hospital Organization Headquarters in Japan (Tokyo, Japan) on November 08, 2019, and Kyoto University School of Medicine (Kyoto, Japan) on August 25, 2023, with approval codes NHO H31-02 and M192. The completion numbers for the authors are AP0000151756, AP0000151757, AP0000151769, and AP000351128. As this research was considered clinical research, consent to participate was required. After briefing regarding the clinical study and approval of the research contents, the participants signed an informed consent form.

## Clinical Research

A multi-center retrospective observational clinical study of subjects who underwent cancer genomic medicine at a cancer medical facility in Kyoto, Japan. This study was reviewed and approved by the Central Ethics Review Board of the National Hospital Organization Headquarters in Japan (Tokyo, Japan) on November 18, 2020, and Kyoto University School of Medicine (Kyoto, Japan) on August 24, 2022, with approval codes NHO R4-04 and M237. All participants agreed to take part in the present study. We have obtained Informed Consent Statements from people participating in clinical studies.

## Author Contributions

All authors had full access to the data in the study and take responsibility for the integrity of the data and accuracy of the data analysis. Y.T., T.H., M.O. and H.A.; Research Conduction, T.H., K.A.; Writing-Original Draft, S.T.; Discussion on Biochemical Function, T.H., K.A. and I.K.; Writing-Review & Editing, I.K.; Visualization, T.H. and I.K.; Supervision, T.H. and I.K.; Funding Acquisition.

---

**Table 1.**
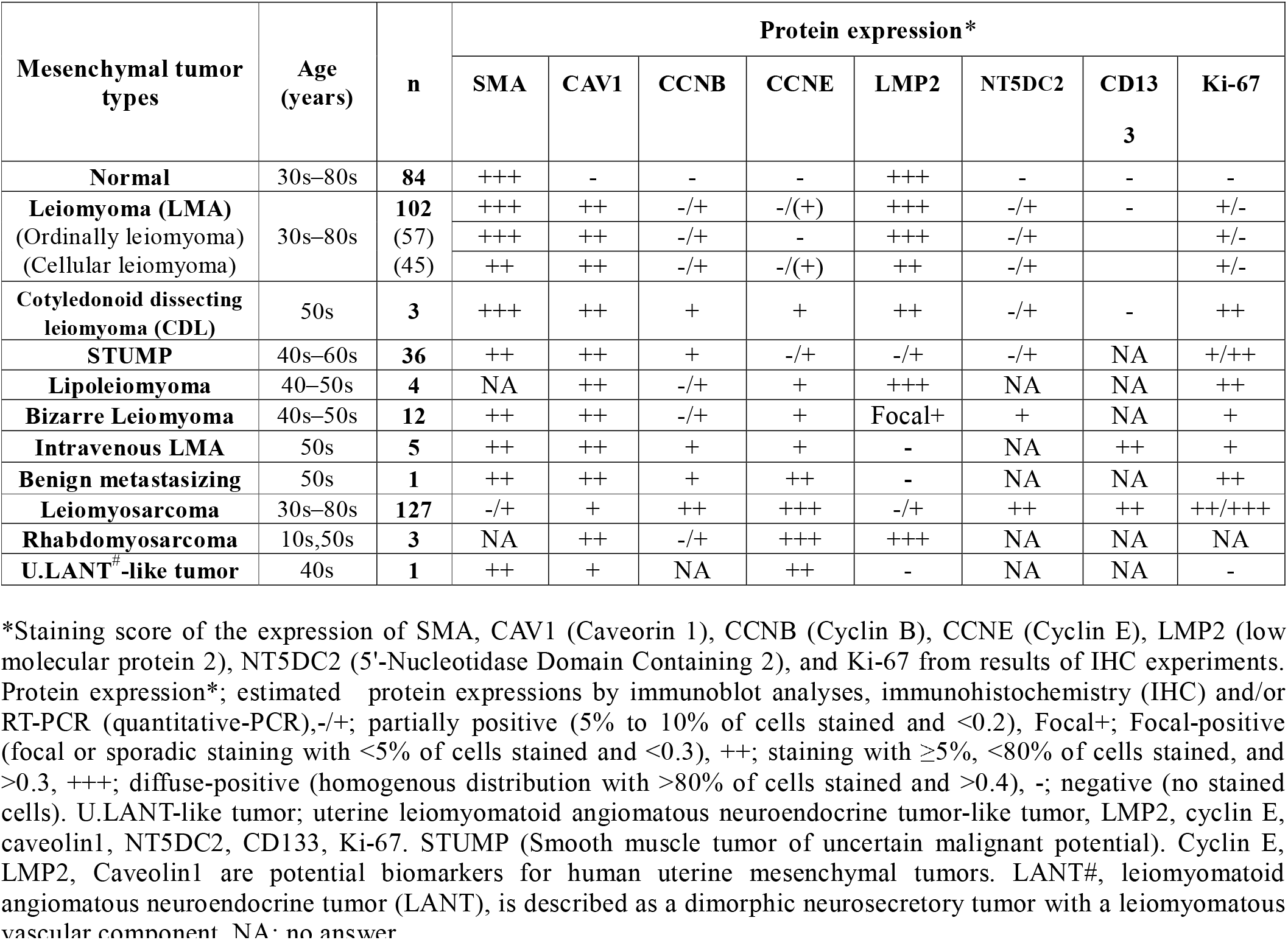
Differential expressions of SMA, Caveolin1, Cyclin B, Cyclin E, LMP2, NT5DC2, CD133, and Ki-67 in human uterine mesenchymal tumors and the uterine LANT-like tumor.

## References

1 Zaloudek C, and Hendrickson MR: Mesenchymal tumors of the uterus, in Kurman RJ (ed): Blaustein’s Pathology of the Female Genital Tract (ed 5). New York, Springer-Verlag, pp561–578, 2002.

2 Gupta AA, Yao X, Verma S, Mackay H, and Hopkins L: the Sarcoma Disease Site Group and the Gynecology Cancer Disease Site Group. Systematic Chemotherapy for Inoperable, Locally Advanced, Recurrent, or Metastatic Uterine Leiomyosarcoma: A Systematic Review. : Clin Oncol (R Coll Radiol). 2013 Jan 5. doi:pii: S0936-6555(12)00362-7.

3 Lin JF, and Slomovitz BM: Uterine sarcoma : Curr Oncol Rep. 10: 512–518, 2008.

4 Amant F, Coosemans A, Debiec-Rychter M, et al: Clinical management of uterine sarcomas. Lancet Oncol. 10: 1188–1198, 2009.

5 Chiang S, and Oliva E: Recent developments in uterine mesenchymal neoplasms. Histopathology. 62:124–37, 2013.

6 Kurma, R.J. Pathology of the Female Genital Tract, 4th ed. New York, Springer-Verlag 4: 499, 2001.

7 Diagnostic Criteria for LMS, Adapted from 2003 WHO Guidelines: World Health Organization

8 Classification of Tumours: Pathology and Genetics, Pathology and Genetics of Tumours of the Breast and Female Genital Organs. 2003; IARC Press, France, 2003.

9 Ravegnini G, Mariño-Enriquez A, Slater J, et al: MED12 mutations in leiomyosarcoma and extrauterine leiomyoma. Mod Pathol. 2012 Dec 7. doi: 10.1038/modpathol.2012.203.

10 Maniatis T: A ubiquitin ligase complex essential for the NF-κB, Wnt/Wingless, and Hedgehog signaling pathways. Genes Dev. 13: 505–510, 1999.

11 Shankaran V, Ikeda H, Bruce AT, et al: IFN-γ and lymphocytes prevent primary tumour development and shape tumour immunogenicity. Nature 410: 1107–1111, 2001.

12 Van Kaer L, Ashton-Rickardt PG, Eichelberger M, et al: Altered peptidase and viral-specific T cell response in LMP2 mutant mice. Immunity 1: 533–541, 1994.

13 Hayashi T, Kodama S, Faustman DL: LMP2 expression and proteasome activity in NOD mice. Nature Medicine 6: 1065–1066, 2000.

14 Hayashi T, and Faustman DL: Development of spontaneous uterine tumors in low molecular mass polypeptide-2 knockout mice. Cancer Res. 62: 24–27, 2002.

15 Hayashi T, Kobayashi Y, Kohsaka S, et al: The mutation in the ATP-binding region of JAK1, identified in human uterine leiomyosarcomas, results in defective interferon-gamma inducibility of TAP1 and LMP2. Oncogene 25: 4016–4026, 2006.

16 Nigg EA: Cyclin-dependent protein kinases: key regulators of the eukaryotic cell cycle. BioEssays 17: 471–480, 1995.

17 Galderisi U, Jori FP, and Giordano A: Cell cycle regulation and neural differentiation. Oncogene 22: 5208–5219, 2003.

18 Orlando DA, Lin CY, Bernard A, et al: Global control of cell-cycle transcription by coupled CDK and network oscillators. Nature 453: 944–947, 2008.

19 Ford HL, and Pardee AB: Cancer and the cell cycle. J. Cell. Biochem. Suppl 32-33: 166–172, 1999.

20 Zhou XY, Wang X, Hu B, Guan J, Iliakis G, and Wang Y: An ATM-independent S-phase checkpoint response involves CHK1 pathway. Cancer Res. 62: 1598–1603, 2002.

21 O’Connor DS, Wall NR, Porter AC, and Altieri DC: A p34(cdc2) survival checkpoint in cancer. Cancer Cell 2: 43–54, 2002.

22 Agarwal R, Gonzalez-Angulo AM, Myhre S, et al: Integrative analysis of cyclin protein levels identifies cyclin b1 as a classifier and predictor of outcomes in breast cancer. Clin. Cancer Res. 15: 3654–3662, 2009.

23 Hayashi T, Yaegashi N, Konishi I. Molecular pathological approach of uterine intravenous leiomyomatosis. Ann Transl Med. 2022 Jul;10(13):724.

24 Hayashi T, Horiuchi A, Sano K, et al: Potential role of LMP2 as an anti-oncogenic factor in human uterine leiomyosarcoma: morphological significance of calponin h1. FEBS Letter 586: 1824–1831, 2012.

25 Hayashi T, Horiuchi A, Sano K, Hiraoka N, Kanai Y, Shiozawa T, Tonegawa S, Konishi I. Mice-lacking LMP2, immuno-proteasome subunit, as an animal model of spontaneous uterine leiomyosarcoma. Protein Cell. 2010 Aug;1(8):711–7.

26 Hayashi, T., et al., Molecular Approach to Uterine Leiomyosarcoma: LMP2-Deficient Mice as an Animal Model of Spontaneous Uterine Leiomyosarcoma. Sarcoma, 2011. p. 476498.

27 Shankaran, V., et al., IFNgamma and lymphocytes prevent primary tumour development and shape tumour immunogenicity. Nature, 2001. 410(6832): p. 1107–11.

28 Delp, K., et al., Functional deficiencies of components of the MHC class I antigen pathway in human tumors of epithelial origin. Bone Marrow Transplant, 2000. 25 Suppl 2: p. S88–95.

29 Nakajima, C., et al., A role of interferon-gamma (IFN-gamma) in tumor immunity: T cells with the capacity to reject tumor cells are generated but fail to migrate to tumor sites in IFN-gamma-deficient mice. Cancer Res, 2001. 61(8): p. 3399–405.

30 Brucet, M., et al., Regulation of murine Tap1 and Lmp2 genes in macrophages by interferon gamma is mediated by STAT1 and IRF-1. Genes Immun, 2004. 5(1): p. 26–35.

31 Chatterjee-Kishore M, Wright KL, Ting JP, Stark GR How Stat1 mediates constitutive gene expression: a complex of unphosphorylated Stat1 and IRF1 supports transcription of the LMP2 gene.. EMBO J. 2000 Aug 1;19(15):4111–22.

32 Nishikawa S, Hayashi T, Uzaki T, Yaegashi N, Abiko K, Konishi I. POTENTIAL LIFE PROGNOSTIC MARKER FOR MESENCHYMAL TUMOR RESEMBLING UTERINE LEIOMYOSARCOMA. Georgian Med News. 2023 Oct;(343):119–126.

33 Zhai YL, Kobayashi Y, Mori A, et al: Expression of steroid receptors, Ki-67, and p53 in uterine leiomyosarcomas. Int J Gynecol Pathol. 18: 20–28, 1999.

34 Akhan SE, Yavuz E, Tecer A, et al: The expression of Ki-67, p53, estrogen and progesterone receptors affecting survival in uterine leiomyosarcomas. A clinicopathologic study. Gynecol Oncol. 99: 36–42, 2005.

35 Wang HX, Wang HM, Li QL, et al: Expression of Proteasome Subunits Low Molecular Mass Polypeptide (LMP) 2 and LMP7 in the Endometrium and Placenta of Rhesus Monkey (Macaca mulatta) During Early Pregnancy. Biology Reprod. 71: 1317–1324, 2004.

36 Yang L, Fang D, Chen H, Lu Y, Dong Z, Ding HF, Jing Q, Su SB, Huang S. Cyclin-dependent kinase 2 is an ideal target for ovary tumors with elevated cyclin E1 expression. Oncotarget. 2015 Aug 28;6(25):20801–12.

37 Moberg KH, Bell DW, Wahrer DC, Haber DA, Hariharan IK. Archipelago regulates Cyclin E levels in Drosophila and is mutated in human cancer cell lines. Nature. 2001 Sep 20;413(6853):311–6.

38 Raso MG, Barrientos Toro E, Evans K, Rizvi Y, Lazcano R, Akcakanat A, Sini P, Trapani F, Madlener EJ, Waldmeier L, Lazar A, Meric-Bernstam F. Heterogeneous Profile of ROR1 Protein Expression across Tumor Types. Cancers (Basel). 2024 May 15;16(10):1874.

39 Hu B, Zhou S, Hu X, Zhang H, Lan X, Li M, Wang Y, Hu Q. NT5DC2 promotes leiomyosarcoma tumour cell growth via stabilizing unpalmitoylated TEAD4 and generating a positive feedback loop. J Cell Mol Med. 2021 Jul;25(13):5976–5987

